# Optimizing Student Success: Leaders’ Perspectives on Advising Practices in Graduate Health Professions Education Programs

**DOI:** 10.1101/2024.12.02.24318324

**Authors:** Holly S Meyer, Anita Samuel, Jennifer Cleland, Lauren A. Maggio, Anthony R. Artino, Emily Scarlett, Paolo C. Martin

## Abstract

**Phenomenon:** Effective graduate advising is crucial for student success but remains an understudied area, particularly in Health Professions Education (HPE). While the importance of advising is widely recognized, faculty often lack adequate preparation and face competing demands. Current research on advising primarily focuses on other disciplines, neglecting the unique challenges of HPE programs. This study aims to address this gap by exploring leaders’ perspectives on advising practices that optimize student success in HPE graduate programs.

**Approach:** A qualitative interview study using narrative inquiry was conducted to explore leaders’ perspectives on advising practices in graduate HPE programs globally. Purposive sampling was used to recruit program leaders across six World Health Organization regions. Semi-structured interviews were conducted and analyzed using narrative inquiry and developmental advising theory as a framework.

**Findings:** Fifteen HPE program leaders described advising practices focused on academic advancement, personal advancement, and community building. Leaders emphasized the importance of clear communication, goal setting, and student support. While advisors valued building strong relationships with students, challenges arose from balancing professional and personal roles. Findings suggest that a developmental advising approach, incorporating academic, personal, and community-building elements, can enhance student success in HPE graduate programs.

**Insights:** This study underscores the critical role of advisors in graduate HPE programs, revealing that effective advising involves both academic guidance and substantial emotional support. It highlights a gap in the existing literature and advocates for the expansion of developmental advising that addresses not only administrative tasks but also the broader developmental needs of students, such as career advancement and personal growth. The findings suggest that to enhance advising practices, graduate HPE programs should invest in ongoing professional development for advisors, adopt holistic advising models, and allocate resources to support students’ integration into professional communities, thereby preparing them more fully for their future careers.

## Introduction

Leaders of graduate programs have been keenly aware of the importance of graduate advising for decades (Council of Graduate Schools, 1991). Winston et al (1984) identified five key functions of a graduate advisor, including providing reliable information, functioning as a departmental and occupational socializer, serving as a role model, and acting as an advocate. In essence, effective graduate advising goes beyond administrative tasks; when done well, it shapes the academic and personal development of students. Yet, faculty members frequently find themselves thrust into advisory roles without adequate preparation, and advising responsibilities often come in addition to a myriad of other professorial duties. Despite the recognized importance of graduate advising, there is currently a gap in research literature on advising practices that support the advisor-advisee relationship. This work aims to begin filling that gap.

There is also a lack of research on leadership practices that support effective advising in graduate Health Professions Education (HPE) programs. From other disciplines, we can gather clues on how this is done through communicating effectively (Tinto, 1993), developing a personal and professional relationship between advisors and advisees (Smith & Allen, 2006), and acknowledging the distinct challenges that both parties bring to the academic journey (Selke & Wong, 1993). On the student side, individuals pursuing graduate studies are often high-achieving adults (Samuel et al, 2024) accustomed to giving advice rather than receiving it. This dynamic creates a complex interplay of perspectives, as students, driven by self-direction and the pursuit of personal goals, must balance the demands of graduate study with the stresses and responsibilities of adult life. Leadership practices can support this relationship by cultivating a supportive and student-centered environment. To do this, leaders can train, support, and recognize advisors for their role in student success (Kezar, Gehrke, & Lindsay, 2018); however, these actions have yet to be explored in graduate HPE programs.

The need to address these gaps are critical, especially given the rapidly growing field of graduate HPE (Tekian, 2023). Our research aims to uncover leaders’ perspectives on advising practices within HPE graduate programs. Specifically, from the perspective of HPE program leaders, how do advising practices optimize student success in graduate HPE programs?

## Methods

### Study design and participant recruitment

We conducted a qualitative interview study informed by our constructivist orientation. We included HPE programs with a master’s and/or PhD degree option. 108 eligible programs were identified through an extensive website search. The programs spanned across the six global geographic regions of the World Health Organization (WHO): Americas, European, South-East Asian, Western Pacific, African, and Eastern Mediterranean (WHO, n.d.). We purposefully recruited in batches to prioritize a proportional representation of each WHO region. Using purposive sampling,(Patton 2002) we started our recruiting efforts with participants who participated in a previous related research study (Schermerhorn, 2023). Subsequently, we targeted WHO regions where response rates were low proportional to the total number of graduate HPE programs in the region (see Table 1). Three attempts were made to contact programs. Based on our research aim, we collectively determined sufficient data was collected based on the distribution of graduate HPE programs across WHO regions and the richness of data collected (LaDonna et al, 2021). In total, we interviewed 15 participants.

**Table 1.**
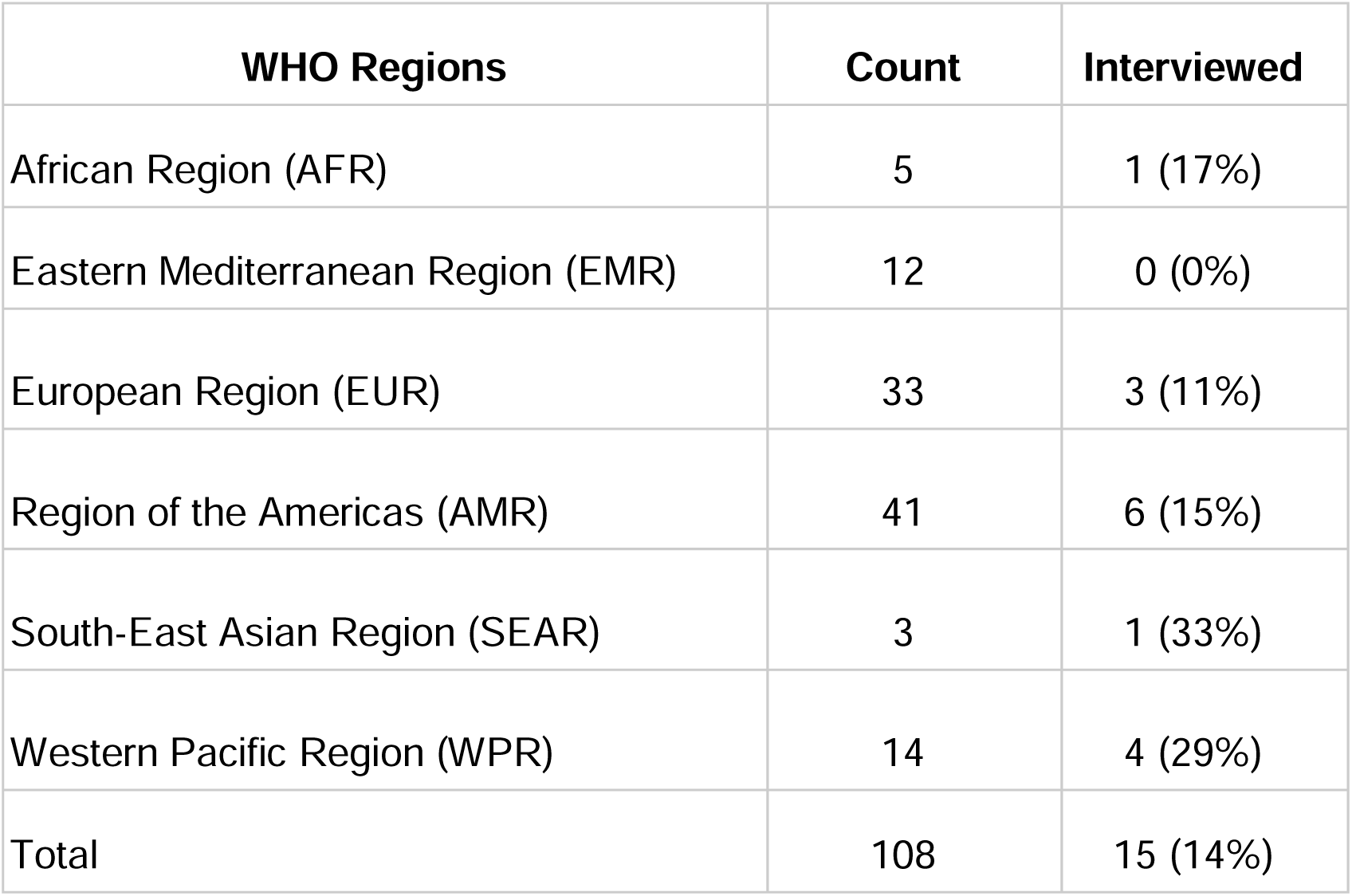
Recruitment of participants across WHO regions.

### Data Collection

We used a semi-structured interview guide to conduct one-on-one interviews. An interview guide addressing the research question was drafted based on the literature and our experience of serving as graduate advisors. The interview guide was piloted, revised, and agreed upon by all authors. The interview questions were drafted to gather comprehensive insights into the advising practices (e.g., the role of the leader; structure and details of the program; advising practices; effectiveness and barriers to advising; and exploration of an ideal advising program). These questions were designed to help us better understand the effectiveness, challenges, and potential improvements in advising practices within graduate HPE programs.

HM and PM conducted all of the interviews in English in the spring of 2023 via video-conferencing. Interviews were approximately 45-75 minutes long and were video recorded. Following the interview, the recordings were transcribed verbatim and the transcripts de-identified. The study was approved by the Institutional Review Board of the Uniformed Services University (Case#: DBS.2022.348).

### Reflexivity

The authors of the present study serve as advisors and leaders within HPE programs; therefore, we see, hear, and interpret participants’ voices within this context. We engaged in an emic approach (Markee 2013), drawing on our insider perspectives and experiences within HPE graduate programs to untangle details of program advising structures. This insider knowledge allowed us to understand the nuances and complexities of advising practices. Simultaneously, we adopted an etic perspective by systematically analyzing and interpreting the data through the lens of developmental advising, with an eye toward ensuring an objective understanding of the advising practices described by the program leaders we interviewed. All authors were experienced researchers in qualitative research methods and relevant theories. Our collective expertise influenced data interpretation.

### Data Analysis

Narrative inquiry was used to study the perspectives of a broad range of individuals who led graduate HPE programs globally. Data from these transcripts were analyzed inductively using initial open-analysis techniques. HM initially coded for advising practices of advisors from four transcripts. HM and PM reviewed this initial set of codes. ES coded the other 11 transcripts and then HM and PM met to refine codes. The remaining members of the research team (HM, LM, AS, ES, AA, JC, PM) were provided with the results of the coded data and sample transcripts for review. The group shared reflections on the data. Through this iterative process, we identified a link between the developmental advising model and the codes present (King, 2005). Via asynchronous email exchanges, consensus of codes and themes was reached.

Developmental advising theory, moving beyond academics to consider learners’ personal, social, and career development (Crookston, 1972), is pertinent to our research given that our study focuses on understanding how advising practices influence student outcomes. Our research aim involves understanding advising practices that extend beyond traditional administrative and course selection requirements of advising to impact student retention, academic performance, and career and personal development. While numerous definitions of developmental advising exist (King, n.d.), we used Creamer and Creamer (1994)’s developmental advising theory as a framework to explore these relationships. which provides a structured, yet adaptable, approach to exploring the complexities and nuances of advising relationships. This theory helped structure our analysis by focusing on key aspects like the advisor-student relationship, the integration of academic and personal development, and the effectiveness of advising strategies. This framework helped us to interpret findings by providing a lens through which to understand how and why certain advising practices impact student outcomes.

## Findings

Fifteen HPE program directors from eight countries and 15 institutions participated in the study. Throughout the interviews, participants discussed both beneficial and ineffective advising practices and the structure of advising in their program. Twelve of the 15 program directors also served as advisors to students within their programs. Although Creamer and Creamer (1994) frame developmental academic advising into six distinct concepts—setting career and life goals, building self-insight and esteem, broadening interests, establishing meaningful interpersonal relationships, clarifying personal values and styles of life, and enhancing critical thinking and reasoning. We reorganized these into the three broader categories of academic advancement, personal advancement, and community building when applied to the context of graduate health professions education advising to emphasize the interconnectedness of these areas in fostering holistic student development and preparing future healthcare professionals. This streamlined framework allowed us to more effectively categorize and analyze the data, highlighting the key areas where developmental advising impacts student growth and success based on leader’s perspectives. To provide context for our findings, we share participant quotes labeled with a letter (e.g., D would indicate the leader expressed the corresponding statement). Table 2 provides a summary of the advising practices from the perspective of graduate HPE leaders.

**Table 2.** Advising practices from the perspective of graduate HPE leaders.

| Advising Practices | Advising practices from the perspective of graduate HPE leaders |
| --- | --- |
| Academic Achievement | • Know and share academic requirements, curriculum changes, and expectations.<br>• Communicate effectively through various channels (group sessions, individual check-ins, emails, etc.).<br>• Manage coursework by mapping out programs of study, selecting courses, and assisting with registration.<br>• Ensure students stay on track with deadlines and progress tracking.<br>• Actively engage students in learning and connecting concepts.<br>• Prepare educators with educational theories and skills.<br>• Guide research projects with feedback and resources.<br>• Refer students to support services and offer additional assistance.<br>• Show genuine interest in students' success.<br>• Encourage professional development and facilitate career opportunities. |
| Personal Achievement | • Serve as a critical friend.<br>• Engage in supportive, non-hierarchical advising.<br>• Collaborate with students, guiding them while allowing ownership of their work.<br>• Facilitate joint knowledge-building.<br>• Draw from personal experience as students.<br>• Serve as a sounding board and provide advice.<br>• Offer insights and feedback based on qualifications.<br>• Help students understand strengths and areas for improvement.<br>• Encourage belief in their capabilities.<br>• Promote new ways of thinking.<br>• Support conflict resolution and personal challenges.<br>• Enable students' journeys and goals. |
|  | ● Provide career guidance and set realistic academic goals.<br>● Offer individualized support.<br>● Address non-program-related issues.<br>● Show genuine interest in students' well-being.<br>● Assist with accommodations and campus familiarization. |
| Community Building | ● Cultivate diversity, inclusivity, and equity.<br>● Promote social justice.<br>● Value all health professionals' voices.<br>● Encourage active participation and peer engagement.<br>● Keep students engaged and motivated.<br>● Highlight the importance of a community of practice.<br>● Create a warm, empathetic, safe, and inclusive environment.<br>● Foster critical thinking and strengths-based narratives.<br>● Build trust and rapport with students.<br>● Model expected behaviors like effective communication, humility, transparency, integrity, fallibility, vulnerability, and accessibility. |

### Academic Advancement

HPE leaders expressed a variety of advising practices used to support student’s academic advancement. These included understanding and communicating academic requirements, curriculum changes, and program expectations. Participants also described helping students manage their coursework by mapping out their program of study, selecting appropriate courses, and assisting with registration. For example, one participant noted: “The purpose is quite explicit: to provide guidance for students to navigate the program, which is highly individualized and self-directed.” (Leader K)

Leaders reported that advisors supported academic advancement through multiple modalities including communication, deadlines, and portfolio checks. Advisors used clear and effective communication including group advising sessions, individual check-ins, and emails. Advisors ensured students stayed on track and made progress toward their goals, sometimes, by using deadlines to create a sense of urgency and structure as needed. Programs that used portfolios noted that their advisors helped to document the student’s process via portfolios. Advisors also used practices that directly help students in their studies by helping them connect concepts across courses or contexts; and providing knowledge of educational concepts, theories, and skills. Leader H expands on this when stating:

> Our goal is often to surface what their educational needs are, articulate them with them and then send them in that direction. Maybe sometimes it’s actually directly teaching them about whatever that might be, but often it’s actually directing them in the… providing resources for them to identify the answers, fulfill the knowledge gap or skills gap themselves.

Some leaders noted that advisors play a role in the students’ research project; helping guide the student along it to make it academically rigorous. Such assistance takes the form of shaping research advisory teams, guiding the selection of research topics, providing feedback on the project, and equipping them with resources.

> To take the student through the thesis process, from the very beginning of it, of focusing the research question and putting together a pre-proposal and then a proposal, taking them through their proposal hearing, helping them with IRB, advising during the data collection and analysis, reviewing and critiquing the final thesis write up, taking them through the thesis defense, all of that.” (Leader M)

Advisors use a variety of strategies to support students more broadly in their education. This includes helping them to transition to higher-level studies (e.g., a master’s student decided to enroll in a PhD program), encouraging them to make connections or seize opportunities, and sponsoring their educational development. Leader G emphasizes this transition by explaining “I’ve had some MA students who are now doing a PhD with me in HPE.” Another leader described connecting with the larger community through networking, “we (the program) have got connections.… so we can enable our students to achieve whatever they want to achieve, by connecting them with other people” (Leader F). When advisors cannot directly support students, advisors sometimes refer students to other supports such as mental wellness programs, financial support, or writing centers. Of note, a small number of programs did not offer any kind of formalized career discussions, “Because not all our participants [students] want to do that…They don’t really care about a career because it’s not a career that they’re building in med ed. They just happen to be doing some teaching because their workplace has said they had to” (Leader J).

### Personal Advancement

In graduate HPE, advising practices supported students in their personal advancement. Advisors emphasized the importance of tailored, empathetic advising practices coupled with clear, effective communication to foster active engagement and a strong advisor-advisee relationship. This support extended beyond academics by offering “non-academic support, more mentally supporting students, emotionally supporting them, and helping them manage their workload, study load, work-life balance, family, work, study, life balance” (Leader E). Participant H said, “it’s important that we know our students and that there’s a strong emotional component.” They support this relationship by engaging in practices including facilitating knowledge building together, serving as a critical friend, being more supportive and less hierarchical, and being collaborative while still giving students ownership of their work. They were able to partner beside the student by acting as a sounding board, offering insight based on their own qualifications or experiences in the program and providing feedback when needed. Advisors can “learn from students, there’s a lot of co-learning that happens. It also maintains my relationships with clinicians, which are important for my program of research. So a lot of the MA students that I supervise in HPE are clinicians whom I do research with” (Leader G).

Many advising practices revolve around student goals. HPE advisors helped promote goal setting, enable the student’s journey, provide opportunities for career discussions, manage professional expectations, assist with a student’s personal goals, and adapt to support students based on their unique needs, goals, and interests. Sometimes enabling the student’s journey took on an advocacy and championing tone,

> “university policies and procedures…most of the time they’re an irritant…You have a good student, they’ll go through and they’ll put lots of barriers in their way, as often happens. So a lot of the time I spend advocating for students, whether it’s for admissions or they’re not moving forward well, but there’s a reason, give them a break type of thing” (Leader C).

Other practices revolved around developing the students and their insight by promoting their understanding of their strengths and areas for improvement, turning weaknesses into learning opportunities, encouraging students to believe in their capabilities, promoting new ways of thinking, helping them with conflict resolution, and helping them work through personal challenges.

Advisors also had to engage in some more assorted practices to assist the student’s personal advancement. This included dealing with non-program-related issues, providing guidance on accommodations and campus familiarization, showing genuine interest in student’s well-being, and also being aware of the challenges of distance learning. Much like developmental academic advising, HPE advisors helped support the growth of the student in a holistic manner, with practices that encourage a close, less hierarchical relationship where they could also learn alongside the student.

However, some programs supported advising without any of the closeness, “I never saw it as a mentor or coaching role. Because I don’t think I had the level of intimacy in the relationship” (Leader J). Sometimes the context of the graduate HPE program prevents this closeness because advisees come from established, professional careers. One leader stated,

> “[I] had a student who had some really, really significant mitigating circumstances and we tried to get him to go to see the pastoral tutor [advisor]. He wouldn’t see him because he said, this person knows me in my professional context and I don’t want him to know that I have this going on in my life. And so it was, it actually mattered quite a lot who this person was at that point, it wasn’t sort of a neutral thing” (Leader O).

### Community Building

HPE leaders also discussed advisor practices that revolved around building an inclusive supportive community that values diversity, equity and inclusion (DEI), belonging, mutual respect, and active participation. Advisors accomplished this by “try[ing] to look at diversity, inclusivity, and equity… whether you are considering those sorts of thrust areas in the research work that you’re doing, in terms of the way we mentor, the way we supervise” (Leader N) or promoting social justice, and ensuring voices of all health professionals are valued.

> [Advisors] balance creating those opportunities for them to participate in the discipline, in the community, while at the same time giving them enough space to try out their ideas, to push against hegemonic thinking, because we tend to control our disciplines very often, we have ways of doing and ways of thinking and ways of writing, that can be quite alien and foreign to a lot of our students…I think the supervisor’s role is to balance those and to enable the journey. (Leader L)

Advisors also created empathic, inclusive environments from the beginning, fostered critical thinking skills, encouraged strength-based narratives, and highlighted the importance of a community of practice. Using a strengths-based narrative was important given the self-critical lens of students,

> “a lot of times the students feel guilty, they haven’t done enough, or what they’ve done isn’t good enough. They are very self-critical. So part of what I do is try and give them very specific feedback about particular things, but also kind of build up their confidence, that they can keep going, that they are enough…You can’t have successful students or graduates if they don’t feel like they are capable” (Leader I).

HPE advisors served as role models, “I model the behavior that I want to see” (Leader A). Additionally, much like developmental academic advising emphasizes the shared nature of the advising relationship, HPE advisors encouraged active participation, motivation, peer engagement, and helped students choose topics they found personally meaningful. “[It’s] also about maintaining their motivation throughout the coursework. So more emotional motivational support maybe, and ensuring their well-being” (Leader E). In HPE, this encouragement may manifest in the selection of a research topic, “advise them always to do something in their research that they’re excited about. It has to be something important to them” (Leader H).

## Discussion

This study identified a range of leaders’ perceptions of advisor-advisee practices within graduate HPE programs. Effective advising was found to encompass both academic and personal support, such as assisting with course selection and providing substantial emotional support. The advisor’s role in creating a supportive community and learning environment was also crucial in helping students develop a sense of belonging and mutual respect. The key findings focused on the necessity for advisors to have up-to-date programmatic knowledge, provide individualized support, and prepare students for their professional careers (Gordon, Habley, & Grites, 2008; Lowenstein, 2005).

### Expanding Developmental Advising in Graduate HPE Programs

This study builds on existing literature by highlighting the need for developmental advising in graduate HPE programs. While previous research has emphasized administrative aspects of advising, such as course selection, this study underscores the importance of addressing the broader developmental needs of students, including career advancement and professional development (Astin, 1984; O’Banion, 1972). The findings align with existing literature on the significance of advisors possessing a deep understanding of program requirements and providing personalized guidance, but they also extend this knowledge by emphasizing the importance of holistic support that fosters self-awareness and decision-making skills (Crookston, 1972; King, 2005).

### Enhancing Advisor Development and Holistic Support in Graduate HPE Programs

The findings suggest that graduate HPE programs should prioritize ongoing professional development and training for advisors to ensure they remain current with program requirements and policies (Habley & McCauley, 2009; Lowenstein, 2005). This is necessary for delivering individualized support and preparing students for their professional careers. Programs might consider adopting apprenticeship models of advising that emphasize mentorship and hands-on learning experiences (Brown & Daly, 2004). Additionally, there is a need to revise policies to support holistic advising practices that attend to the whole student, not just administrative tasks. Developmental advising offers an approach that promotes a diverse and inclusive environment, fostering student engagement and growth (Winston & Polkosnik, 1984). Future research should explore how program leaders can establish clear expectations for advising, develop effective training methodologies, and implement systems to recognize and promote advisors (CASEL, 2023; Gordon, Habley, & Grites, 2008). Understanding the role of institutional support, particularly in the context of tenure and promotion, could provide valuable insights into incentivizing effective advising practices (Pascarella & Terenzini, 2005; Ryan & Deci, 2017; Tinto, 1993).

Program leaders should prioritize ongoing professional development and training for advisors to ensure they remain informed about program requirements and policies (Habley & McCauley, 2009; Lowenstein, 2005). They must also promote individualized and holistic advising practices that cater to each student’s unique needs and support their overall well-being (Gordon, Habley, & Grites, 2008). Additionally, leaders should allocate resources to help connect students with professional communities and ensure that advisors are equipped to prepare students for their future careers (Kezar, Gehrke, & Lindsay, 2018).

### Strengths and Weaknesses of the Study

Our focus on the perspectives of HPE program leaders, a crucial yet often overlooked stakeholder group in advising practices. However, the study’s reliance on English-language interviews may have excluded non-English-speaking participants, limiting the diversity of perspectives. Additionally, the study focused on master and PhD degree programs, and findings may not be fully representative of advising practices in other types of programs, such certificate programs.

## Conclusion

Findings from this work emphasize the critical role of advisors in providing both academic and personal support within graduate HPE programs. The results further highlight the need for up-to-date programmatic knowledge and individualized guidance. This work underscores the importance of expanding developmental advising to address students’ broader career and personal development needs, a practice that remains under-explored in this context. To achieve this, program leaders should prioritize ongoing professional development for advisors, ensure holistic advising practices, and allocate resources to connect students with professional communities, thereby better preparing them for their future careers.

## Data Availability

All data produced in the present study are available upon reasonable request to the authors.

