## Appendix 1 for "Optimizing Student Success: Leaders’ Perspectives on Advising Practices in Graduate Health Professions Education Programs"

Protocol #: HPEAdvising

### Interview Guide:
### Leaders of Health Professions Education Programs (HPE)

**Research purpose**

The aim of this study is to identify coaching, mentoring, and advising practices that optimize student success in health professions education (HPE) graduate programs. We believe that strong HPE programs have goals for their students and that mentorship, advising, and coaching practices can help HPE programs reach the program goals and student's goals. Therefore, when we identify and match coaching, mentoring, and advising practices to specific HPE program goals we optimize the program's success and learner's success.

**Overview**

This interview is designed for leaders (e.g., directors, deputy directors, hiring managers) of health profession programs.

**How to use this guide**

*This interview guide is organized into a framework of 5 groups of questions. For each grouping, the objective for the category is listed, followed by guiding questions to meet the objective.*

*Text written in italics is guidance for the interviewer.*

*We want to interviews to use the language of the program. Therefore, we ask questions to probe how the program names faculty-student relationships (e.g., advising/mentoring/coaching). Once the program names the term the interviewer will use that term.*

*At the beginning of the session, please introduce yourself and relay the following information:*

The purpose of this interview is to uncover the faculty-student practices where learners can obtain advice, counseling, and direction in their training. My role, as the interviewer, is to facilitate your opportunity to share your insights, I am not here to judge, criticize or validate your thoughts and opinions.

This is a safe space; your opinions and your words will not be associated with your identity or likeness; likewise, the identity of your home institution or program will never be tied to your comments. This session is recorded, but only to facilitate an accurate transcript of what we discuss today. The only persons who will have access to these recordings will be the research team for the purpose of transcription. The final transcript will be de-identified. Quotes of your words may appear in a publication but will be anonymized. Research material such as the de-identified transcripts may be accessed by other researchers and reviewers to validate the accuracy of the data. The discussion we have today is confidential, however, I cannot ensure confidentiality if you disclose any illegal activity, intentions to harm yourself or others.

It is anticipated that this interview will take about 60 minutes. If at any time you wish to stop, just let me know and the interview will be over. Last step before we begin – with your permission, I will begin recording by using the Google Meet record function which will capture our video and audio feed. I will also turn on my audio recorder as a backup. All recordings will only be accessible by myself and the research team and stored on a secure system. Once the transcription is complete and verified these recordings will be destroyed. May I have your consent to participate in the study and your consent to begin recording?

> *If the participant declines to give consent for the recording, the participant will be offered to reschedule to have a second investigator join the call as a scribe.*

—-

This interview consists of five parts. First, I ask some general questions about you and your role; second, about the program, third, about success in HPE programs; forth, about facult-student relationships in your program; and finally, a summary with next steps.

**Section 1: About you**
Let's start with the 1st section to learn more about you and your role.

1. What is your role?
2. What are your responsibilities?
3. How long have you been with your institution and in this role?
4. How did you end up in this role?

**Section 2: About the program**
Let's shift gears and talk about your program.

1. What year did the program start?

2. Why was it established?
3. What certificates and/or degrees does your program offer?
4. What is required for each certificate/degree?
    a. *Probe to learn more about* # of courses, research requirements, etc for each certificate and degree?
5. How many students graduate each year? What degrees?
6. How many students are currently enrolled?
7. How many full-time faculty?
8. How many staff members?
9. What format is your program – online/blended/in-person?
    a. Can you tell me more about waht that means for your program?
    b. How much time is spent face-to-face vs online?
10. How much time is required to complete the program?
    a. *Probe for each certificate/degree.*
11. Are faculty local or online?
12. Can you share with me some general information about your student body including percentage of your currently enrolled learners who identify as a person of color? Average age range? Specialities of students?
13. Is feedback from students on the program collected? If so, how?
14. If a student is struggling in the program, how are they identified?
    a. What actions are taken?
15. What role do alumni play in your program?

**Seciton 3: Defining success in HPE programs**

We're moving onto the third section of our interview. I would like to better understand how your pogram defines success.

16. What makes a successful HPE graduate in your program?
17. How do you know if an HPE graduate is successful?
18. Can you share any general information about what the programs look for in applicants? For example, what makes a successful applicant to your program?
19. If a student has a question about how to successfully complete the program who would they contact?
    a. *If an example is needed, say,* "Questions might include number of courses to take or suggestions for best practices on balancing resarch and clinic demands."
    b. *If it depends on the student's question, ask* "Can you tell me more about the different responsiblities of staff and faculty?"

c. *If students is directed to a specific individual, ask* "What does your program call this person's role?"
    i. Probing question: Do faculty or staff members have responsibility for providing students with advise or direction while in the program?
    ii. Probing question: The higher education langage describes programs having student-faculty relationships often called coaching, mentoring, advising or other terms. Does this resonate with your program?

**Section 4: Student-Faculty Relationships**

20. The next series of questions is to better understand advising/mentoring/coaching *(use the term the participants used in the previous question)* in your program. Can you describe how advising/mentoring/coaching works in your program?
    a. *Probe to better understand:*
        i. *Alignment*. How and when are (advisor/mentors/coaches/etc) aligned with students?
            1. Are students aligned with more than one advisor/mentor/coach?
        ii. *%*. Roughly what percentage of your (faculty/staff) serve in this role?
        iii. *Ratio*. What is the adviee/advisor ratio? Is there even distribution of students across faculty/staff? How is this determination made?
        iv. *Qualifications*. What qualifications do (advisors/mentors/coaches/etc) need to serve in this role?
        v. *Cert vs degree*. Does advising/mentoring/coaching change depending on a learner being a certificate vs degree learner?
        vi. *Academic vs degree*. Does advising/mentoring/coaching vary based on the learner's stage in the program - for example being in course work vs in research?
        vii. *Purpose*. What is the purpose (advising/mentoring/coaching) in your program?
        viii. *Responsibilities*. What are the responsibilities of the (advisors/mentors/coaches/etc)?
        ix. Interactions. What interactions do advisors/mentors/coaches have with students?
            1. Which of these are required by the program vs at the faculty/staff or student's discretion?
            2. At what points during the year or program do the intereactions occur? (e.g. every semester, end of every year, after assessments, etc)

x. *Training*. What training do faculty receive to serve in this role?
xi. Workload. Does (advising) count toward faculty's other responsibilities, like teaching load?
xii. *Other responsibilities*. Do (advisors/mentors/coaches) have other responsibilities besides (adivsing/mentoring/coaching)?
*xiii*. *Effectiveness*. How do you know if an advisors/mentor/coach is effective? Can you give me an example?
xiv. *Characteristics*. What characteristics make an effective (advisor/mentor/coach)?

21. What characteristics make an effective advisee/mentee?
22. What is your role in advising/coaching/mentoring in your program?
23. Has advising/mentoring/coaching at your program looked this way for the past five years or have changes occurred to this system?
    a. If changes, probe to better understand these changes.
24. Can you tell me a story of a successful *advising/mentoring/coaching* relationship in your program?
25. What contributes to a successful advising/mentoring/coaching relationship in your program?
26. I am hearing that (*list characteristics described in the previous question*) contribute to success. Did I capture this correctly?
27. I would like to better understand each of these. What activities or actions contribute to building ____ (fill in with contribution)?
    a. Repeat the question for each characteristic.
28. Can you tell me a story of an unsuccessful advising/mentoring/coaching relationship in your program?
29. What contributes to advising/mentoring/coaching not being successful or not as successful as it could be?
30. I am hearing that (list characteristics described in the previous question, eg distrust, etc) contribute to the challenges. Did I capture this correctly?
31. What suggestions do you have for rectifying these challenges?
32. (*If applicable*) How do online vs in-person interactions impact advising/mentoring/coaching in your program?
33. If the skies were blue, and time and resources were not limited, what would the ideal advising/mentoring/coaching interactions look like in your program? Why?

**Section 5: Summary Questions**

In this last question set, I would like to ask you a few summary questions.

1. Before we end the interview, I want to make sure you had a chance to share all of your thoughts and opinions. Is there something we didn't talk about that you think is important?
2. Is there anything else you would like to add?

3. Would you be willing to be contacted by our research team for further clarification on your answers?
4. This study is part of a larger body of research. While we are currently only focusing on HPE leaders, the next phase of the study is to discuss these questions with faculty members who serve in an advising/mentoring capacity and learners/alumni in the program. Would you be willing to share some names of faculty and students we could contact to interview?

On behalf of the research team, I'd like to thank you for participating in this interview.

Thank you for participating in this study – your thoughts today have been invaluable. I am going to go ahead and stop the recording. If you have any questions comments or concerns, feel free to reach out to me or one of the other team members. Our contact information is in the original email that you received from us. Can I answer any questions now before we disconnect?

*A note to the interviewer: please remember to turn off the recording functions of the Google Meet and the backup recorder. Ensure that the files are securely stored in the designated location. Lastly, take a few moments to reflect on this interview session.*
